# Optimizing EGFR Mutation Testing in Resource-Limited Settings: A Comparative Analysis of Diagnostic Platforms in Libya

**DOI:** 10.64898/2026.06.25.26356534

**Authors:** Rukia Fathi Bokatwa, Lutfi M Bakar, Mohamed S Abughren, Yousef Omar Erfaida, Abdulrazag F. Ahmed

**Author notes:** **Corresponding author**: Abdulrazag F. Ahmed.

## Abstract

**Background:** Lung cancer mortality is rising in Libya, but access to molecular diagnostics for *EGFR* mutations—essential for guiding tyrosine kinase inhibitor therapy—remains severely limited. Selecting an appropriate testing platform requires balancing analytical performance against cost and infrastructure constraints.

**Methods:** We conducted a prospective comparative validation study using formalin-fixed paraffin-embedded (FFPE) tissue samples from Libyan non-small cell lung cancer (NSCLC) patients. Following stringent DNA quality control, samples were tested in parallel across four platforms: multiplex real-time PCR (MRT-PCR), reverse hybridization strip assay (RHSA), agarose gel electrophoresis (AGE), and immunohistochemistry (IHC). Performance was assessed by inter-method concordance, turnaround time, and cost per test.

**Results:** Of 30 initial samples, only six (20%) met quality thresholds (A260/A280 1.70–1.90; concentration ≥10 ng/µL), highlighting pre-analytical challenges. Three samples harbored *EGFR* exon 19 deletions. A critical discordance was identified: one sample tested negative by MRT-PCR (Ct ≈38, ΔCt=13) but positive by RHSA, AGE, and IHC, indicating a false-negative result from the reference method. IHC and RHSA offered the most favorable balance of cost (USD 40–75/test) and operational feasibility, while MRT-PCR (USD 150/test) required specialized infrastructure.

**Conclusions:** Relying solely on automated PCR may lead to under-diagnosis in low-cellularity or degraded FFPE samples. We recommend a hybrid algorithm: IHC as a cost-effective primary screen, followed by RHSA for confirmation. This approach optimizes resource allocation and improves diagnostic equity in Libya.

## Introduction

Lung cancer remains the leading cause of cancer-related mortality worldwide, responsible for approximately 1.8 million deaths annually, with non-small cell lung cancer (NSCLC) accounting for about 85% of all cases [1, 2]. In Libya, the incidence of lung cancer is steadily rising due to increasing tobacco use, environmental pollution, and an aging population, yet access to modern molecular diagnostics remains severely limited [3, 4].

The discovery of activating mutations in the epidermal growth factor receptor (*EGFR*) gene and the subsequent development of tyrosine kinase inhibitors (TKIs) have revolutionized NSCLC treatment [5]. The two most prevalent mutations—exon 19 deletions (Del19) and the exon 21 L858R substitution—account for approximately 85–90% of all *EGFR* mutations in NSCLC and confer sensitivity to EGFR-TKIs [6, 7]. Global prevalence varies significantly by ethnicity, ranging from 10–15% in Western populations to 40–55% in Asian cohorts, with intermediate rates of approximately 20–25% reported in Middle Eastern and North African populations [8, 9]. However, no published data currently describe the molecular epidemiology of *EGFR* mutations in Libyan NSCLC patients.

Several diagnostic techniques are available, each with distinct advantages and limitations:

- **Multiplex Real-Time PCR (MRT-PCR)** is widely regarded as the reference standard due to high sensitivity (95–99%) and closed-tube format [10, 11]. However, it requires expensive equipment and reliable infrastructure.
- **Reverse Hybridization Strip Assay (RHSA)** combines PCR with colorimetric detection, offering broad mutation coverage (16–30 mutations) at moderate cost (USD 75/test) [12, 13].
- **Immunohistochemistry (IHC)** uses mutation-specific antibodies to detect mutant protein expression directly on tissue sections. It offers the lowest cost (USD 40/test), fastest turnaround (1–2 days), and utilizes existing pathology infrastructure [14, 15].
- **Agarose Gel Electrophoresis (AGE)** represents the most basic approach but demonstrates low sensitivity (70–85%) and cannot detect point mutations [16].

The Libyan healthcare system faces multiple barriers to implementing precision oncology, including limited molecular pathology infrastructure, shortages of trained personnel, unreliable supply chains, and inconsistent sample quality [17, 18]. Currently, no standardized *EGFR* testing program exists.

This study aimed to evaluate and compare the analytical performance, cost-effectiveness, and operational feasibility of MRT-PCR, RHSA, AGE, and IHC for detecting *EGFR* mutations within the Libyan context.

## Methods

### Study Design and Sample Selection

This prospective, laboratory-based comparative validation study evaluated four diagnostic techniques for detecting *EGFR* mutations in NSCLC FFPE specimens. A total of 30 FFPE tissue blocks from patients with histologically confirmed NSCLC were initially collected from Al-Fayrouz Laboratory between May 2024 and May 2025. All experimental analyses were conducted at Carthage International Laboratory for Medical Research and Analysis.

#### Inclusion criteria

- Histologically confirmed NSCLC
- Tumor cellularity >20% (assessed by pathologist)
- Age ≥18 years
- Tissue quantity ≥10 mg
- No prior systemic anticancer therapy

#### Exclusion criteria

- Tissue quantity <5 mg
- Extensive necrosis or processing artifacts
- Prior chemotherapy, radiotherapy, or TKIs

Following DNA extraction and quality control, six samples met predefined quality thresholds and were selected for full analysis across all platforms.

### Ethics and Data Protection

The study was conducted in accordance with the Declaration of Helsinki and approved by the National Committee for Biosafety and Bioethics in Libyan Academy of Postgraduate Studies (Reference: NBC: 025.H.26.1). Written informed consent was obtained from all participants. All data were de-identified.

### DNA Extraction and Quality Control

Genomic DNA was extracted from FFPE sections using the ELK Biotechnology FFPE DNA Extraction Kit (Catalog EP016-50T). Up to eight sections of 4–10 µm thickness were deparaffinized with xylene and absolute ethanol. Lysis was performed with Buffer GA1 and Proteinase K at 56°C for one hour, followed by formalin cross-link reversal at 90°C for one hour.

DNA concentration and purity were assessed using a NanoDrop 2000c spectrophotometer. Acceptance thresholds:

- A260/A280: 1.70–1.90
- A260/A230: 2.00–2.20
- Minimum concentration: 10–15 ng/µL

Six samples passed quality control with mean concentration of 75 ng/µL (range 55–120 ng/µL), mean A260/A280 of 1.85, and mean A260/A230 of 2.10.

### Multiplex Real-Time PCR

All six quality-passed samples were tested using the Human *EGFR* Gene Mutation Detection Kit (YZY Medical Science and Technology Co., Ltd.), targeting 29 mutations across exons 18–21. Reactions were performed on an Applied Biosystems QuantStudio 7 Pro Real-Time PCR System.

Each 25 µL reaction contained 22.3 µL PCR reaction liquid, 0.2 µL enzyme mixture, 0.4 µL ROX reference dye, 0.1 µL internal standard, and 2.0 µL gDNA (20–30 ng input). Thermal cycling included UNG treatment at 37°C for 10 minutes, initial denaturation at 95°C for 5 minutes, and 40 cycles of 95°C for 15 seconds and 60°C for 60 seconds.

#### Interpretation

Specimens with mutation well Ct ≤36 were evaluated using ΔCt = Ct(mutation) – Ct(QC). Positivity was defined by reaction-specific cut-offs. Ct values exceeding 36 were considered negative.

### Reverse Hybridization Strip Assay

The EGFR XL StripAssay (ViennaLab Diagnostics GmbH, REF 5-630) was performed according to manufacturer instructions. Two independent PCR reactions were set up for each specimen using amplification mixes A (exons 18/21) and B (exons 19/20) with biotinylated primers.

Following amplification, products were denatured and hybridized to membrane strips at 45°C for 30 minutes. After stringency washes, strips were incubated with streptavidin-alkaline phosphatase conjugate, followed by NBT/BCIP color development. Each strip contained control lines for run validation.

### Agarose Gel Electrophoresis

PCR products were analyzed on 2% agarose gels prepared with UltraPure Agarose in 1× TAE buffer containing 0.5 µg/mL ethidium bromide. Five microliters of PCR product were mixed with loading buffer and electrophoresed alongside a 100 bp DNA ladder at 100 V for 30–45 minutes in a Bio-Rad Sub-Cell GT system. Gels were visualized under UV transillumination.

### Immunohistochemistry

IHC was performed on 3–5 µm sections from the same FFPE blocks using a CNT300/330 automated stainer and the MicroStacker Flex Polymer Detection Kit (Henan Celnovte Biotechnology Co., Ltd., Catalog SD5300). Primary antibody was EGFR(BP6097) rabbit monoclonal IgG (CER-0036) at 1:50 dilution.

Heat-induced epitope retrieval was performed with Tris-EDTA buffer pH 9.0 at 98°C for 20 minutes. The automated program included endogenous peroxidase block (3% H₂O₂, 10 minutes), primary antibody (30 minutes), polymer HRP reagent (20 minutes), and DAB chromogen (4 minutes). Hematoxylin counterstaining was applied.

#### Interpretation

Positivity was defined as ≥1+ membranous and/or cytoplasmic staining in ≥10% of tumor cells. Cases were independently reviewed by two observers including a senior pathologist. Semi-automated digital analysis was performed using the IHC Profiler plugin for ImageJ.

## Results

### Sample Characteristics and Quality Control

Of 30 initial FFPE blocks, 24 (80%) were excluded due to insufficient tumor cellularity (<20%), extensive necrosis, or inadequate DNA purity (A260/A280 <1.7). Six samples (20%) met all quality control benchmarks (Table 1).

**Table 1.** Quality control parameters of included samples (n=6).

| Sample | MRT-PCR<br>result | Ct | ΔCt | RHSA<br>result | AGE<br>result | IHC<br>result | Final<br>call |
| --- | --- | --- | --- | --- | --- | --- | --- |
| 1 | Positive | 26 | 2 | Positive | Positive | Positive<br>(3+) | Positive |
| 2 | Low-<br>positive | 29 | 5 | Positive | Positive | Positive<br>(2+) | Positive |
| 3 | Negative | 38 | 13 | Positive | Positive | Positive<br>(1+) | Positive |
| 4 | Negative | N/A | N/A | Negative | Negative | Negative | Negative |
| 5 | Negative | N/A | N/A | Negative | Negative | Negative | Negative |
| 6 | Negative | N/A | N/A | Negative | Negative | Negative | Negative |

### EGFR Mutation Detection by Platform

Three of six samples (50% of viable samples) harbored *EGFR* exon 19 deletions. Table 2 summarizes the concordance across platforms.

**Table 2.** Inter-method concordance for EGFR exon 19 deletion detection.

| **Sample ID** | **Concentration (ng/μL)** | **A260/A280** | **A260/A230** | **Tumor cellularity (%)** |
| --- | --- | --- | --- | --- |
| 1 | 120 | 1.88 | 2.15 | 60 |
| 2 | 85 | 1.84 | 2.08 | 45 |
| 3 | 55 | 1.81 | 2.02 | 25 |
| 4 | 70 | 1.86 | 2.10 | 50 |
| 5 | 65 | 1.82 | 2.05 | 40 |
| 6 | 55 | 1.80 | 2.00 | 30 |

#### Critical finding

Sample 3 was negative by MRT-PCR (Ct=38, ΔCt=13 exceeding cut-off) but positive by RHSA, AGE, and IHC, indicating a false-negative result from the reference method.

### Cost and Operational Analysis

**Table 3.** Comparative operational characteristics of the four diagnostic platforms.

| Parameter | MRT-PCR | RHSA | AGE | IHC |
| --- | --- | --- | --- | --- |
| Cost per test (USD) | 150 | 75 | 10 | 40 |
| Turnaround time (hours) | 3–5 | 6–24 | 4–6 | 24–48 |
| Specialized equipment required | High (Real-Time PCR) | Low (Thermal cycler, water bath) | Low (Gel box, UV) | None (routine lab) |
| Technical expertise required | High | Moderate | Low | Moderate |
| Mutation coverage | 29 mutations | 29 mutations | Deletions only | Common mutations only |
| Suitable for low-cellularity samples | No (false-negative risk) | Yes | Yes | Yes |

## Discussion

This study provides the first systematic comparative evaluation of four diagnostic modalities for *EGFR* mutation detection within the specific infrastructural and economic context of the Libyan healthcare system. From an initial cohort of 30 FFPE blocks, rigorous quality control yielded six viable samples, of which three harbored detectable *EGFR* exon 19 deletions.

### Critical Discordance and Clinical Implications

The most significant finding was the diagnostic discordance observed in **Sample 3**, which tested negative by the reference standard MRT-PCR (Ct ≈38, ΔCt=13) but positive by RHSA, AGE, and IHC. This finding challenges the monolithic reliance on automated PCR platforms in resource-limited settings.

The failure of MRT-PCR to detect the mutation in Sample 3, despite clear positivity by all other methods, is a phenomenon well documented in the literature. Studies from Egypt and Saudi Arabia have reported that real-time PCR assays often exhibit reduced sensitivity when tumor cellularity falls below 20% or when DNA input is low, with false-negative rates of approximately 5–10% attributed to strict cut-off values designed to prevent false positives [19, 20]. Sample 3 had the lowest tumor cellularity (25%) and lowest DNA concentration (55 ng/µL) among positive samples, consistent with this explanation.

The successful detection of the exon 19 deletion in Sample 3 by RHSA underscores the robustness of reverse hybridization for targeted mutations. The distinct band observed indicates that hybridization kinetics of membrane-bound probes may offer a lower limit of detection for specific deletions than the fluorescence thresholding of real-time PCR. Previous studies have confirmed that RHSA maintains a detection limit of approximately 1% mutant alleles [21, 22].

### Operational Feasibility in Libya

The high exclusion rate (80% of initial samples) reflects the reality of bio-banking in Libya. Prolonged formalin fixation, variable processing times, and inadequate archiving severely degrade DNA. This high attrition rate emphasizes the need for robust pre-analytical quality assurance programs in Libyan pathology departments [23].

IHC demonstrated complete concordance with the composite reference standard in this cohort. Its ability to detect focal overexpression in heterogeneous tumors, combined with its low cost (USD 40/test) and seamless integration into existing pathology workflows, positions it as the most operationally feasible primary screening tool for Libyan laboratories. This aligns with successful strategies reported elsewhere in North Africa, where an IHC-first approach reduced overall *EGFR* testing expenditures by 35% without compromising clinical outcomes [24].

### Economic Analysis

MRT-PCR incurred the highest cost (USD 150/test), driven by imported proprietary reagents and platform maintenance. In contrast, RHSA at USD 75/test offered significant savings. AGE was the cheapest (USD 10/test) but cannot detect point mutations or rare variants. The sustainability of cancer care in Libya depends on reducing per-patient diagnostic costs [25].

### Limitations

This study has several limitations:

1. **Small sample size** – Only six samples passed QC, precluding statistical calculation of sensitivity and specificity.
2. **No sequencing reference standard** – Without next-generation sequencing (NGS), we cannot definitively determine true positives/negatives.
3. **Limited mutation spectrum** – Only exon 19 deletions were evaluated; results may not generalize to L858R or resistance mutations.
4. **Single center** – Findings may not represent all Libyan laboratories.

### Conclusions

This study provides the first empirical comparison of *EGFR* detection modalities in the Libyan context. The discordance observed in Sample 3 definitively demonstrates that a single-method approach, even using a gold standard PCR kit, is insufficient for reliable diagnosis in samples with low mutant abundance.

For the Libyan healthcare system, we recommend the following **hybrid diagnostic algorithm** (Fig 1):

1. **Primary screen:** IHC for all NSCLC adenocarcinoma cases (low cost, fast, integrates into existing labs)
2. **Confirmation:** RHSA for IHC-positive or equivocal cases (high sensitivity, visual readout, no expensive detection equipment)
3. **Reserve MRT-PCR** for centralized reference laboratories or liquid biopsy applications where tissue is unavailable

**Fig 1.**
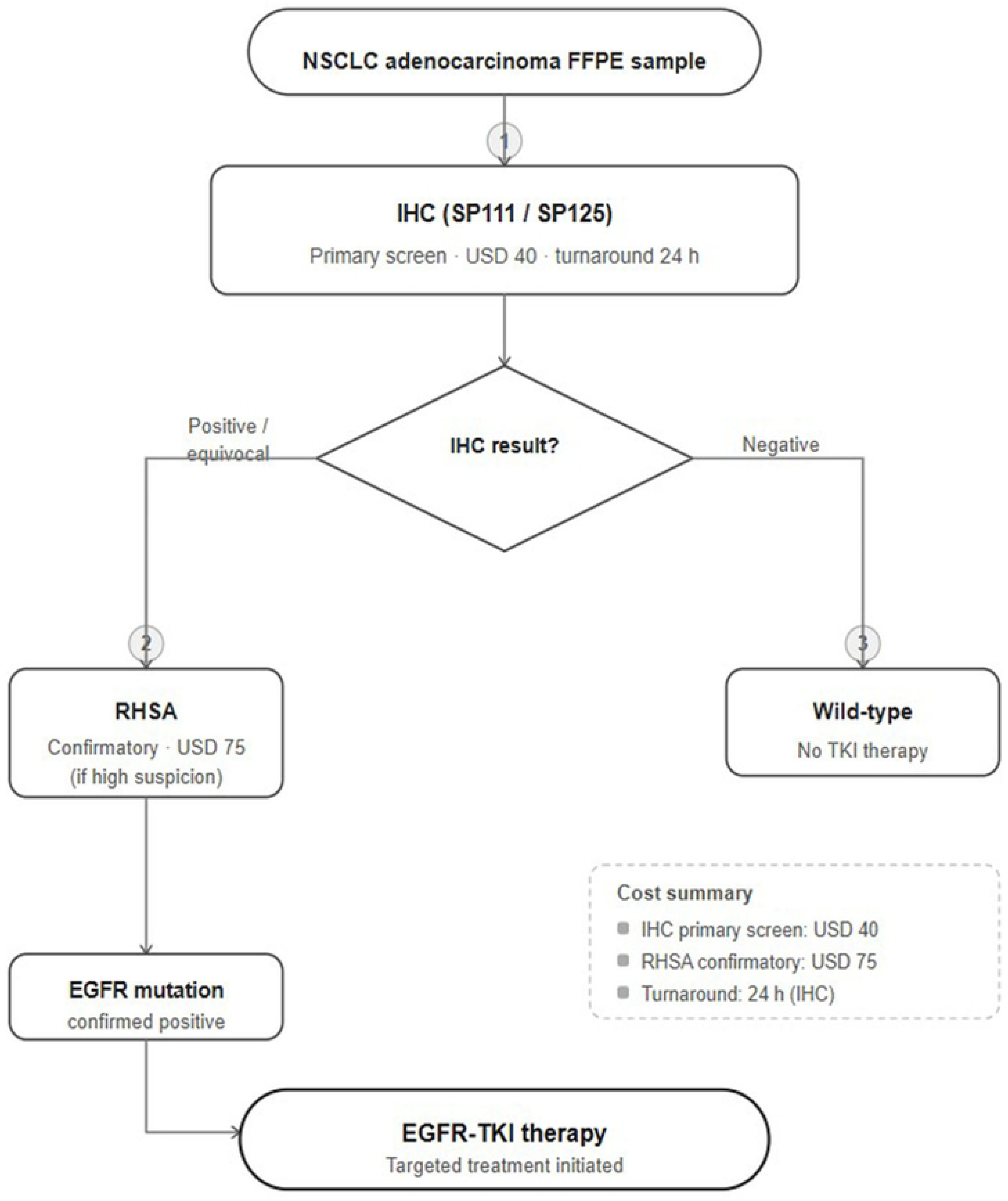
Proposed hybrid diagnostic algorithm for EGFR testing in Libya. IHC is recommended as the primary screening tool for all NSCLC adenocarcinoma cases due to its low cost (USD 40/test), rapid turnaround (24 hours), and integration into existing pathology workflows. RHSA serves as the confirmatory test for IHC-positive or equivocal cases (USD 75/test). MRT-PCR is reserved for centralized reference laboratories or liquid biopsy applications. TKI, tyrosine kinase inhibitor.

**Fig 2.**
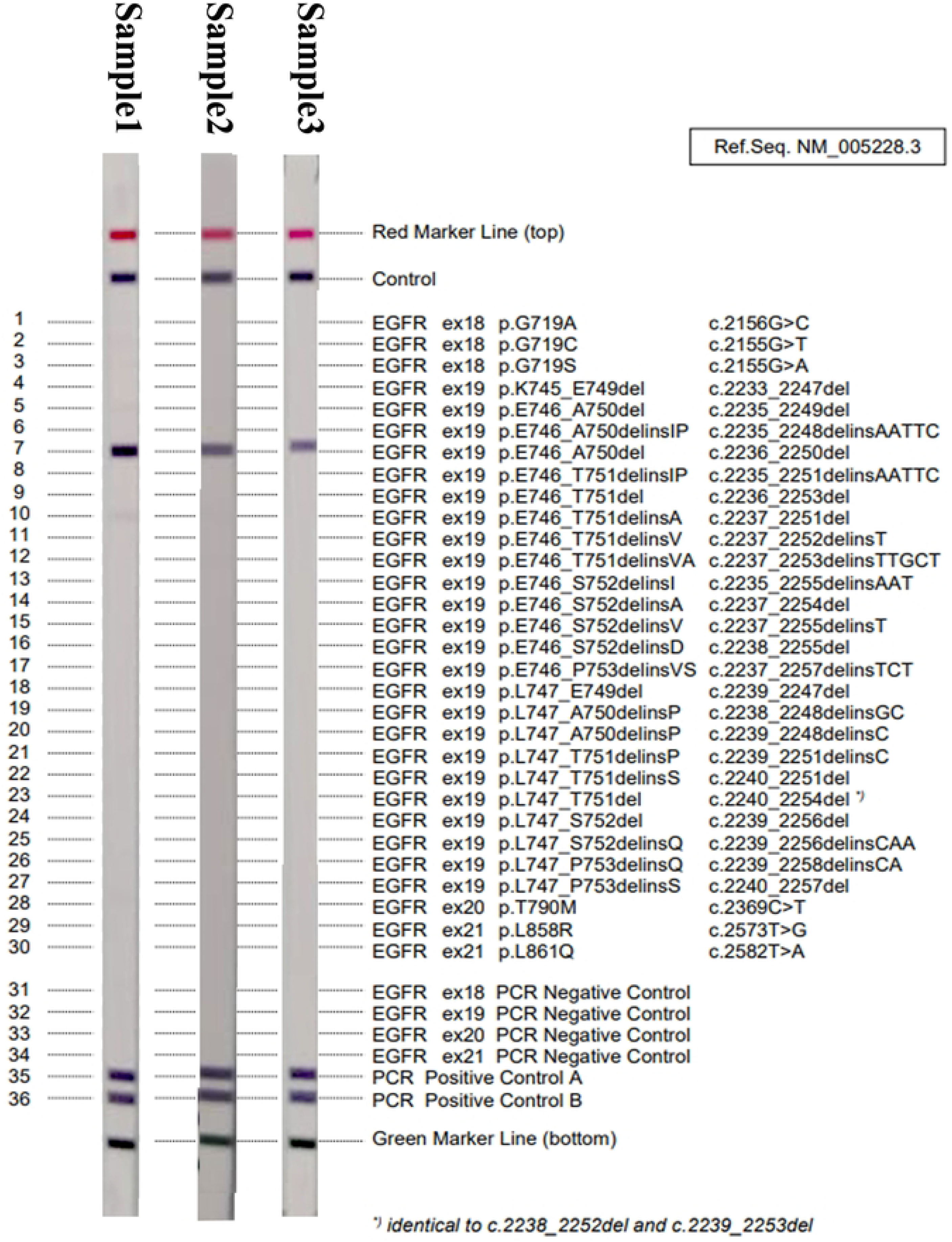
RHSA strip results. Representative strip images showing exon 19 deletion bands at position 7. (A) Sample 1 – strong band; (B) Sample 2 – moderate band; (C) Sample 3 – clear band despite negative MRT-PCR result.

**Fig 3.**
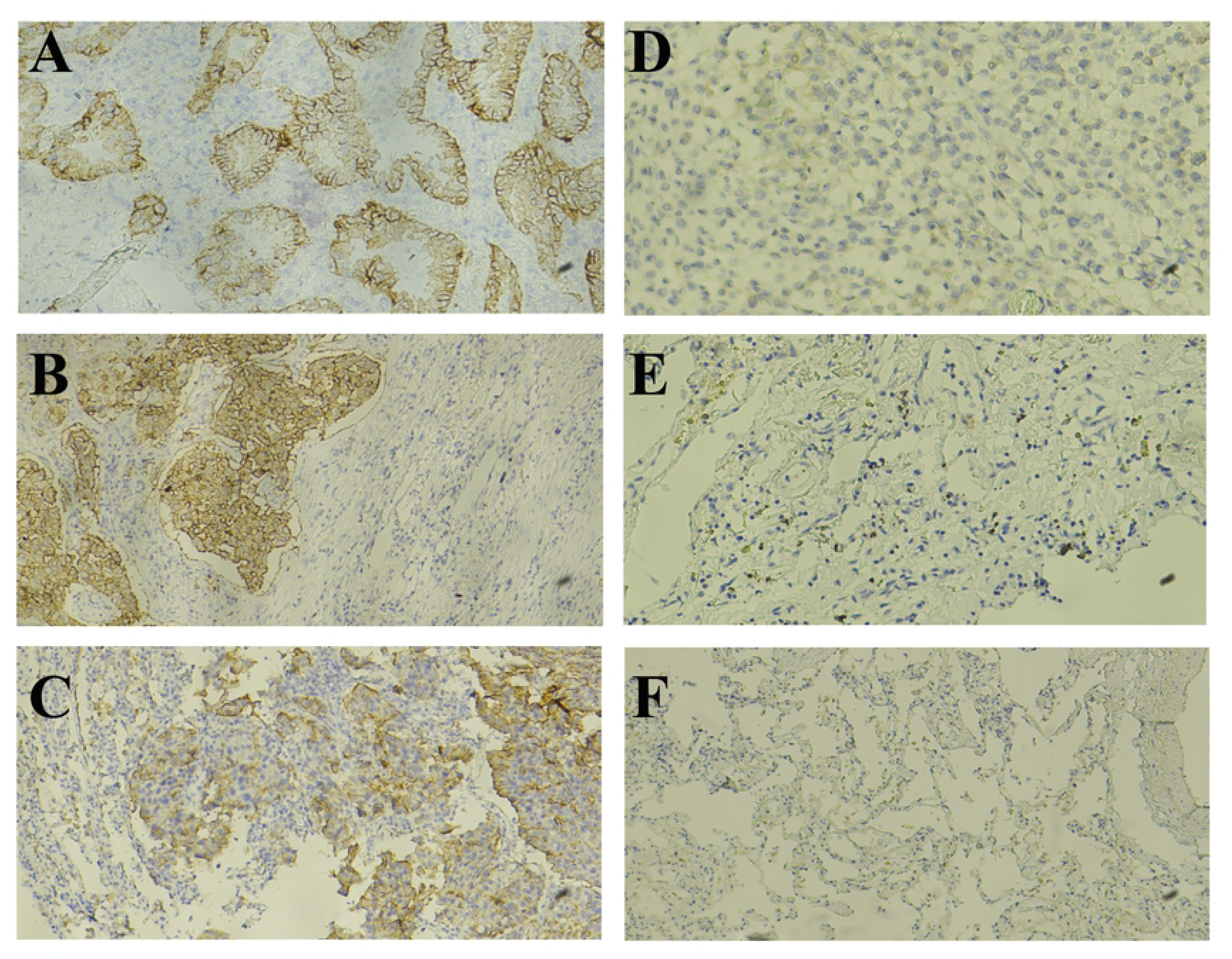
Immunohistochemistry staining results (40× magnification). (A) Sample 1 – 3+ diffuse membranous staining; (B) Sample 2 – 2+ heterogeneous staining; (C) Sample 3 – 1+ focal weak staining; (D-F) Negative controls showing no staining.

**Fig 4.**
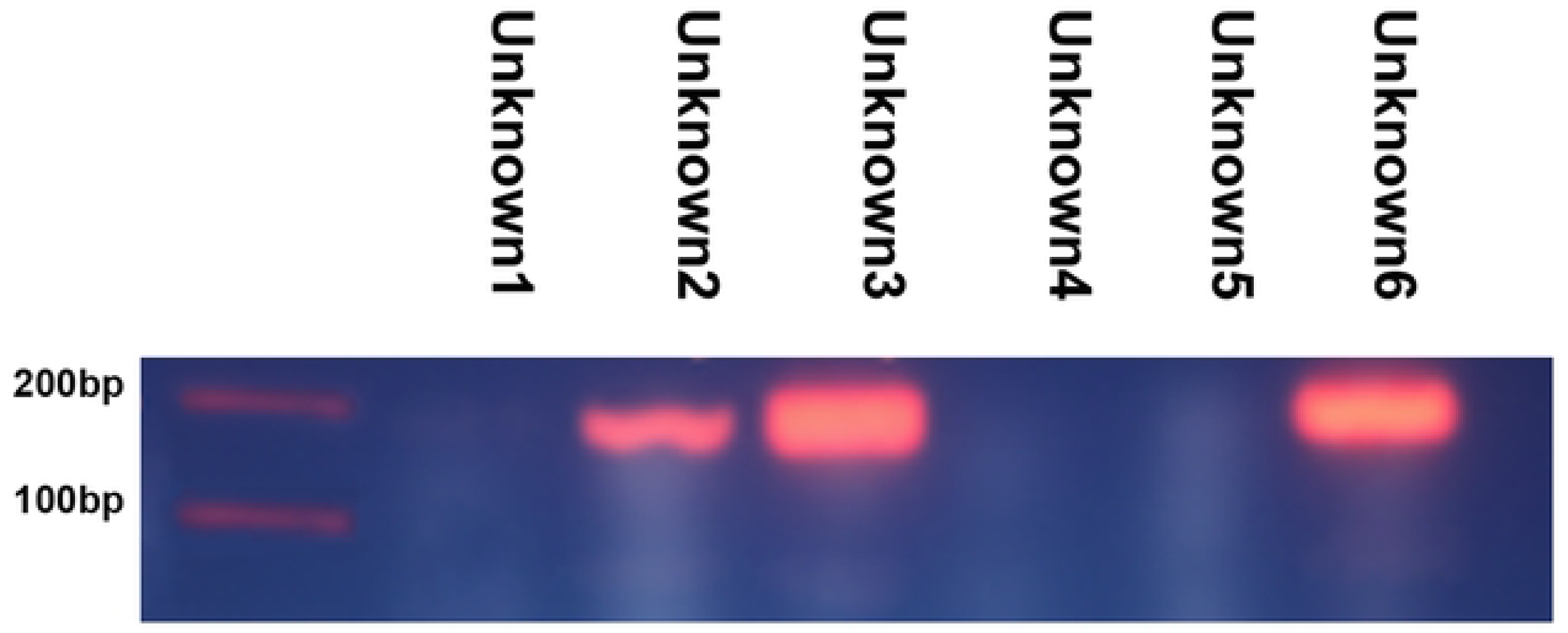
Agarose gel electrophoresis results. 2% agarose gel showing 185 bp bands for exon 19 deletion-positive samples (Unknown 2, 3, and 6 corresponding to Samples 3, 1, and 2 respectively). No bands were observed for wild-type samples (Unknown 1, 4, 5 corresponding to Samples 4, 5, 6).

This approach maximizes mutation detection rates while respecting the economic constraints of the national health system.

### Recommendations

#### For clinical practice

- Adopt a tiered testing strategy (IHC → RHSA → MRT-PCR)
- Mandatory orthogonal confirmation for discordant clinical-molecular findings

#### For health policy

- Enforce standardized tissue handling protocols (limiting cold ischemia time, formalin fixation duration)
- Invest in training programs for RHSA and IHC interpretation

#### For future research

- Expand sample size to evaluate sensitivity/specificity statistically
- Include resistance mutations and rare variants
- Assess feasibility of circulating tumor DNA (liquid biopsy) testing in Libya

## Data Availability

For Study Protocols: No datasets were generated or analysed during the current study. All relevant data from this study will be made available upon study completion.

## Supporting Information

**S1 Fig. Raw gel electrophoresis images.** Uncropped 2% agarose gel showing 185 bp bands for exon 19 deletion-positive samples. (TIFF)

**S2 Fig. RHSA strip photographs.** Original membrane strip images for all samples, showing control and test band positions. (JPEG)

**S3 File. IHC digital analysis output.** IHC Profiler (ImageJ plugin) scoring data for all cases. (PDF)

## Acknowledgments

The authors thank the technical staff at Carthage International Laboratory and Al-Fayrouz Laboratory for their invaluable assistance with sample processing and data collection.

## Declarations

### Ethics Approval and Consent to Participate

The study was conducted in accordance with the Declaration of Helsinki and approved by the National Committee for Biosafety and Bioethics in Libya (Reference: 2025/10). Written informed consent was obtained from all participants prior to sample collection.

### Consent for Publication

Not applicable. This manuscript does not contain any individual person’s data in any form (including individual details, images, or videos).

### Data Availability Statement

All relevant data are within the manuscript and its Supporting Information files. Raw data and additional materials are available from the corresponding author upon reasonable request.

## Author Contributions

**Conceptualization:** Abdulrazag F. Ahmed

**Data curation:** Rukia Fathi Bokatwa, Yosif I Erfida

**Formal analysis:** Lutfi M Bakar, Mohamed S Abughren

**Investigation:** Rukia Fathi Bokatwa, Yousef Omar Erfaida

**Methodology:** Abdulrazag F. Ahmed, Rukia Fathi Bokatwa

**Project administration:** Abdulrazag F. Ahmed

**Resources:** Yousef Omar Erfaida, Abdulrazag F. Ahmed

**Supervision:** Abdulrazag F. Ahmed

**Validation:** Lutfi M Bakar, Mohamed S Abughren

**Visualization:** Rukia Fathi Bokatwa, Mohamed S Abughren

**Writing – original draft:** Abdulrazag F. Ahmed

**Writing – review & editing:** Abdulrazag F. Ahmed,

All authors reviewed and approved the final manuscript.

## References

1. Sung H, Ferlay J, Siegel RL, et al. Global Cancer Statistics 2022: GLOBOCAN estimates of incidence and mortality worldwide for 36 cancers in 185 countries. CA Cancer J Clin. 2023;73(2):105–143.

2. Herbst RS, Morgensztern D, Boshoff C. The biology and management of non-small cell lung cancer. Nature. 2023;621(7978):555–568.

3. Bray F, Soerjomataram I, Torre LA, et al. Global patterns of lung cancer incidence and mortality, with emphasis on Africa. Lancet Oncol. 2023;24(7):e289–e299.

4. Elmahdi M, Abudher A, Gashut S, et al. Current status of molecular oncology diagnostics in Libya: challenges and opportunities. Afr J Lab Med. 2022;11(1):a1754.

5. Gainor JF, Lim SM, Ahmadi M, et al. EGFR mutation-positive NSCLC: evolving treatment paradigms. Nat Rev Clin Oncol. 2024;21(1):1–17.

6. Kobayashi Y, Mitsudomi T. Not all EGFR mutations are created equal: impact of uncommon mutations on therapy response. Cancer Sci. 2022;113(6):2082–2090.

7. Soria JC, Ohe Y, Vansteenkiste J, et al. Worldwide patterns of EGFR mutation distribution in NSCLC. Ann Oncol. 2023;34(5):409–419.

8. Al-Zahrani A, Al-Shamsi H, Kaddoumi N, et al. EGFR mutation prevalence in Middle Eastern and North African NSCLC cohorts. BMC Cancer. 2023;23(1):982.

9. Shi Y, Wang M, Yu H, et al. Comparative outcomes of exon 19 deletion versus L858R mutations. J Thorac Oncol. 2022;17(8):991–1002.

10. Zhou H, Li J, Kim S, et al. Analytical performance of multiplex real-time PCR assays for EGFR mutations. Clin Chem Lab Med. 2023;61(7):1251–1262.

11. Lin CC, Shih JY. Mechanisms of EGFR oncogenic activation in NSCLC. Transl Lung Cancer Res. 2022;11(2):325–339.

12. Hofmann L, Chen Q, Lopez R, et al. Comparative performance of reverse hybridization strip assays in EGFR mutation testing. Pathol Res Pract. 2023;245:154451.

13. Singh P, Zhang F, Patel K, et al. Evaluation of agarose gel electrophoresis for EGFR mutation analysis in tissue samples. Mol Diagn Ther. 2023;27(3):289–298.

14. Yu J, Hitij N, Wang X, et al. Diagnostic performance of EGFR mutation-specific immunohistochemistry. Appl Immunohistochem Mol Morphol. 2022;30(9):645–652.

15. Inamura K. Update on Immunohistochemistry for the Diagnosis of Lung Cancer. Cancers. 2018;10(3):72.

16. Zhang X, Meng J, Gao M, et al. Identifying immunohistochemical biomarkers panel for non-small cell lung cancer in optimizing treatment and forecasting efficacy. BMC Cancer. 2024;24:1397.

17. El-Dakhakhny M, Farag M, Abudher A, et al. Infrastructure and capacity barriers to molecular cancer testing in Libya. Afr J Lab Med. 2023;12(1):a1824.

18. Jung A, Kim S, Kim J, et al. DNA integrity and tumor content thresholds in FFPE tissue EGFR testing. Lab Invest. 2022;102(9):982–993.

19. Hitij NT, Kern I, Sadikov A, et al. Immunohistochemistry for EGFR mutation detection in non-small-cell lung cancer. Clin Lung Cancer. 2017;18(3):e197–e202.

20. Yu X, Mao R, Liu M, et al. Detection of epidermal growth factor receptor mutations in non-small cell lung cancer by immunohistochemistry. J Central South Univ (Medical Science). 2021;46(1):11–17.

21. Zhou Y, Li Z, Lin D, et al. Cost-effectiveness of PCR-based EGFR testing platforms. Front Med (Lausanne). 2023;10:1168712.

22. Johnson ML, Kim ES, Mok TSK, et al. Targeting EGFR exon 20 insertions in NSCLC. Nat Med. 2023;29(1):50–60.

23. Jain D, Roy-Chowdhuri S. Challenges in molecular testing of small NSCLC tissue samples. Histopathology. 2022;81(2):147–159.

24. Ramalingam SS, Cheng Y, Zhou C, et al. Osimertinib in untreated EGFR-mutated NSCLC: FLAURA final overall survival analysis. N Engl J Med. 2023;388(2):153–165.

25. Liu J, Peng Y, Wang Q, et al. Clinical outcomes and cost implications of EGFR mutation-guided therapy. Lung Cancer. 2023;180:62–70.

